# Short-term frailty index fluctuations in older adults: Noise or signal?

**DOI:** 10.1101/2024.10.09.24315145

**Authors:** Erwin Stolz, Anna Schultz, Emiel O. Hoogendijk, Olga Theou, Kenneth Rockwood

## Abstract

**Background:** Reversible short-term fluctuations in the frailty index (FI) are often thought of as representing only noise or error. Here, we assess (1) size and source of short-term FI fluctuations, (2) variation across socio-demographics, (3) association with chronic diseases, (4) correlation with age, frailty level, frailty change, and mortality, and (5) whether fluctuations reflect discrete health transitions.

**Methods:** Nationwide, biweekly longitudinal data from 426 community-dwelling older adults (70+) were collected in the FRequent health Assessment In Later life (FRAIL70+) study using a measurement burst design (5,122 repeated observations, median of 13 repeated observations per person). We calculated the intraindividual standard deviation (iSD) of the FI and used location-scale mixed regression models.

**Results:** Mean iSD was 0.04 (SD=0.03). Fluctuations were driven foremost by cognitive problems, somatic symptoms, and limitations in instrumental and mobility-related activities of daily living. Short-term fluctuations correlated with higher FI levels (r=0.62), one-year FI change (r=0.26), and older age (+3% per year). Older adults who took to bed due to a health problem (+50%), those who had an overnight hospital stay (+50%), and those who died during follow-up (+44%) exhibited more FI fluctuations.

**Conclusions:** Short-term FI fluctuations were neither small nor random. Instead, as older adults become frailer, their measured health also becomes more unstable; hence short-term fluctuations in overall health status can be seen as a concomitant phenomenon of the aging process. Researchers and clinicians should be aware of existence of reversible fluctuations in the FI over weeks and months and its consequences for frailty monitoring.

## INTRODUCTION

Frailty in older adults results from a cumulative decline in multiple physiological systems and is defined^1^ as a state of increased vulnerability toward negative outcomes^2–6^ such as falls, disability, hospitalization, and death when exposed to (minor) internal or external stressors. Used as a tool for risk stratification, frailty assessment allows for more patient-centred care, which in turn can result in better outcomes and the avoidance of harm for older adults.^7,8^ The cumulative deficit model^9^ is one of two main operationalizations^7,10,11^ and depicts frailty as a state variable characterising older adults’ overall health^12,13^ based on a large number (30+) of age-related health deficits^12–14^, including symptoms, diseases, functional impairments, disabilities, and abnormal measurements, summarised in a continuous frailty index (FI). The FI is calculated as the proportion of accumulated health deficits^15^, ranging from 0 to 0.7 ^16–18^.

How the FI changes throughout old age has been studied for more than a decade^19^, with a focus on population-level (average) long-term changes or trajectories and associated risk factors. It has been repeatedly shown, for example, that the FI increases with age^20–23^ and that women^23,24^ and older adults with a low socioeconomic position^25–27^ tend to have a higher average FI. Whereas such mean group differences are fairly well-established, much less is known about individual-level FI trajectories. Where mean FI differences and changes seem to follow regular patterns (e.g., 4-6% increase per year of life^21,28^), individual FI trajectories have been described as more stochastic and irregular, involving sequences of both improvements *and* declines^29^ via many small changes but also some big jumps^30^. In this context, it is important to conceptually differentiate^31^ between enduring long-term FI changes (intraindividual change) and reversible short-term FI fluctuations (*intraindividual variability*), the latter which can be thought of as vertical oscillations around individuals’ long-term FI trajectory. Empirical quantification of FI fluctuations was first provided using cross-national data of community-dwelling older adults in Europe.^32^ The authors reported FI fluctuations to amount to 0.04 and 0.05 on average, which was considerable against the average FI of 0.11 (men) and 0.16 (women). However, that study was based on sparse biannual health survey data, which may underestimate FI fluctuations, as many health changes among older adults likely go unnoticed over such a long timeframe. More recently, indirect evidence on short-term FI fluctuations was provided by two methodological studies^33,34^. Using longitudinal data over three months both studies reported a substantial standard error of measurement (SEM) of 0.05-0.06. Reversible FI fluctuations within individuals over a short period of time – as reflected by the SEM – are usually considered – explicitly in methodological studies and implicitly in most substantive work – to represent only random error or noise.

Building on a long tradition of substantive research on intraindividual variability^35–37^, we argue that reversible short-term within-person fluctuations in older adults’ overall health status (the FI) could also contain a signal, that is, systematic information about the aging process. Several arguments suggest that short-term FI fluctuations could be informative: (1) FI fluctuations could reflect a string of event-related meaningful discrete health transitions over weeks and months. For example, when a high-functioning older adult experiences a serious fall injury and breaks an arm, which together with a longer-than-necessary hospital stay results in difficulties with basic care for him- or herself over several weeks before recovering almost but not fully in the subsequent months.^38–40^ (2) Short-term FI fluctuations could also originate from several chronic illnesses characterised by fluctuating symptomatology such as pain in osteoathritis^41^, lung function in chronic lung disease^42^, negative affect in depression^43^, or neuropsychiatric symptoms in dementia^44^. Short-term fluctuations have also been reported more generally with regard to pain^45^ and sleep^46^. (3) Short-term FI fluctuations may reflect age-related inconsistency in physical and cognitive functioning. For example, fluctuations over days, weeks, and months in disability^47^ as well as cognition^48,49^ tend to increase with chronological age.

Knowledge about FI fluctuations is also important practically: if short-term fluctuations are substantial and vary across older adults, then established between-person group differences and aggregate-level estimates of what constitutes a clinically meaningful change^50,51^ would have limited meaning and use in clinical practice.

Here, we aim to extend our understanding of the nature of short-term fluctuations in health status (FI) among older adults by leveraging newly collected longitudinal data. Our goal is to assess whether short-term FI fluctuations indeed represent only error and noise, or whether they exhibit systematic properties. Specifically, we evaluate (1) the size and source of short-term FI fluctuations, whether FI fluctuations (2) are associated with specific diseases, (3) vary across sociodemographic characteristics, (4) increase with age, FI level, FI change, and are associated with mortality risk, and (5) reflect discrete health events.

## METHODS

### Data

For this analysis, longitudinal data were collected in the FRequent health Assessment In Later life (FRAIL70+) study, where a survey agency recruited a nation-wide sample of community-dwelling older adults in Austria (Supplementary Methods 1). Data were collected under a measurement burst design^31,52,53^ which allows to simultaneously assess both short-term FI fluctuations (within-burst) and long-term change (between bursts). In the first burst, 426 participants aged 70 years and above were interviewed biweekly up to 7 times (mean run time=87 days), starting in September 2021. Within the first burst, retention rates ranged between 98.3% and 95.3%. The biweekly assessment schedule was adopted because previous research^54^ suggested that acute changes in the FI over two weeks are informative. One year after the end of the first burst, a second burst was conducted with 378 returning participants (between burst retention rate = 88.7%), again including up to 7 repeated biweekly interviews (mean run time=76 days) (Supplementary Figure 1). Within the second burst, retention rates were also high (94.2%-76.7%), except for the very last interview (53.5%). The first interview in each burst was conducted in person, follow-up interviews were conducted by phone. For a small sub-sample (n=40), all interviews of the first burst were conducted in person to assess effects of the interview mode and to obtain repeated physical performance tests. The study was approved by the Ethics Committee of the Medical University of Graz (EK-number: 33-243 ex 20/21 1035-2021).

### Variables

The calculation of the FI at each time point followed standard procedure^15,55^ and used 40 self-reported health deficits (Supplementary Table 1). These included chronic diseases, limitations in basic, instrumental, and mobility-related activities of daily living, somatic symptoms, depressed affect, sensory impairment, physical inactivity, poor self-rated health, and (test-based) impaired cognition. Health deficits generally referred to the last two weeks. The FI was calculated by dividing the sum of the 0-1 coded health deficits by the number of health deficits.

Additional variables included time-constant socio-demographics (sex: men/women; age in years; level of education: low/medium/high; living alone: no/yes; social support (3-item Oslo Social Support Scale): low/medium/high); time-varying health events during the last two weeks (taking to bed due to health problem: no/yes; a fall: no/yes; an overnight hospital stay: no/yes); assessment number within burst (1-7); interview mode (personal/telephone); burst (first/second); 1-year mortality (no/yes, obtained via proxy-interviews or contacting local municipality). Missing data in the covariates were minimal: 1% (n=5) in the social support scale and 0.2% (n=2) for 1-year mortality), both of which were addressed with a random forest imputation procedure (out-of-bag error = 0.03).

### Statistical analysis

First, we quantified the magnitude of short-term FI fluctuations by calculating the intraindividual standard deviation (iSD), that is, how much older adults’ FI varied within persons across repeated biweekly assessments. To assess where short-term FI fluctuations originate, we calculated within-burst intra-individual variability also at the health deficit level using the intraindividual interquartile range (iIQR). To assess whether fluctuations are driven by ‘unreliable’ self-reports, as a sensitivity analysis, we calculated iIQRs also for three health deficits based on objective physical performance measures (gait speed, chair rise test, grip strength, see Supplementary Methods 2) for the sub-sample (n=40) which was assessed in person throughout the first burst. Second, to assess whether short-term FI fluctuations varied by socio-demographic characteristics, chronological age, the person-specific average FI level, one-year FI change, and mortality status, as well as acute health events, we used location-scale mixed regression models^56,57^. These models extend the standard mixed regression approach by allowing to not only model the mean (‘location’) but also the within-person variation (‘scale’) as a second outcome. Coupled with the measurement burst design, this approach allows to separate (1) FI level (between-person, across burst), (2) FI change, (within-person, between burst), and (3) FI fluctuations (within-person, within-burst), assess their interrelationship (random effect correlations), and the effects of predictor variables there on. Specifically, we started with an unconditional means location-only model (M0) to assess the size of variance components. This was followed by an unconditional location-only growth curve model (M1) adding burst and within-burst assessment as time variables. The latter was added to ensure that within-burst repeated measurements are fully de-trended. This was then compared with an unconditional location-scale growth curve model (M2) where the scale parameter was added as a second outcome to assess whether explicit modelling of FI fluctuations improved model fit. Then, we included time-stable predictors and as well as death between bursts (M3) for both outcomes. Finally, to assess whether FI fluctuations reflect discrete health events (bedrest, falls, hospital stays), we also included these as time-varying predictors in the final location-scale model (M4). For more details on the statistical approach, see Supplementary Methods 3.

## RESULTS

### Participants

Of 426 participants, 64.6% were women, 66.0% lived alone, and mean age was 77.7 (SD=5.4, range=70-97) years. 19.3% of the participants had a low (compulsory education), 54.2% a medium (vocational training), and 26.5% a high (high school and above) level of education. 46.3% had strong, 42.0% moderate, and only 11.6% poor social support. 50.9% of the participants reported one or more episodes of bedrest, 25.4% one or more falls (with or without subsequent injury), and 8.9% hospital stays during the two three-month measurement bursts. The 426 participants provided 5,123 FI measurements, that is, a median 13 repeated observations per person.

### Descriptive data

The mean FI at baseline was 0.19 (SD=0.14) and the empirical sub-maximum (99^th^ percentile) was 0.61. Other characteristics of the FI included a right-skewed distribution, a positive association with chronological age, higher FI values among women than men, higher values among those with lower education, and considerably higher values among those few (n=11) who died between bursts (Supplementary Figure 2). Descriptive statistics of the FI per biweekly assessment are shown in Supplementary Figure 3, which indicate no discernible population-level trend within the two 3-month bursts. The intraindividual mean (iMean), i.e., the average FI per person per burst (over up to seven repeated assessments) was 0.18 (SD=0.13) in the first and 0.19 (SD=0.13) in the second burst, i.e., a 5.5% annual increase. This was even the case although those 48 participants who did not return for the second burst had a considerably higher average iMean of 0.26 (SD=0.20) in the first burst.

### Main Results

The mean iSD was 0.04 (SD=0.03, lower quartile=0.01, median=0.03, upper quartile=0.05, range=0.00-0.22) in both bursts, i.e., amounted to 1.6 health deficits. This is non-negligible if compared against the average of about 7 health deficits. FI fluctuations were also clearly visible (Figure 1) at the individual-level: While many older adults exhibited only minor fluctuations (iSD, grey background) across their iMean (dashed line), some exhibited large jumps, both up and down. In the last row (first tile left), for example, one person’s FI changed repeatedly between 0.35 (14 deficits) and 0.50 (20 deficits) before dropping to 0.25 (10 deficits), all over 3-months’ time. Short-term FI variability amounted to 29% (=0.04/0.14) of the between-person FI variance at baseline. Older adults’ iSD showed highly similar characteristics (Figure 2) to the FI itself, notably a weak correlation with chronological age and a strong positive correlation with the iMean (Figure 3, plot A and B), i.e., more frail older adults tended to have more fluctuations. Also, Figure 3 (plot C) shows that the magnitude of one-year FI changes was often within the range of short-term FI fluctuations. The association between older adults’ frailty level (iMean) and their short-term fluctuations (iSD) also showed when looked at from a categorical perspective: Among those who were robust (iMean≤0.25, n=334), only 24% had unstable health (iSD>0.04). Conversely, among those who were frail^58^ (iMean>0.25, n=91), 84% were unstable. One consequence of short-term FI fluctuations is that the classification (robust vs. frail) becomes inconsistent for a number of cases: During the seven measurements of the first burst, for example, 30.5% of older adults would be considered “frail” (FI≥0.25) at one biweekly assessment but were “robust” (FI<0.25) at another, that is, their FI fluctuated one or multiple times across the cut-off.

**Figure 1:**
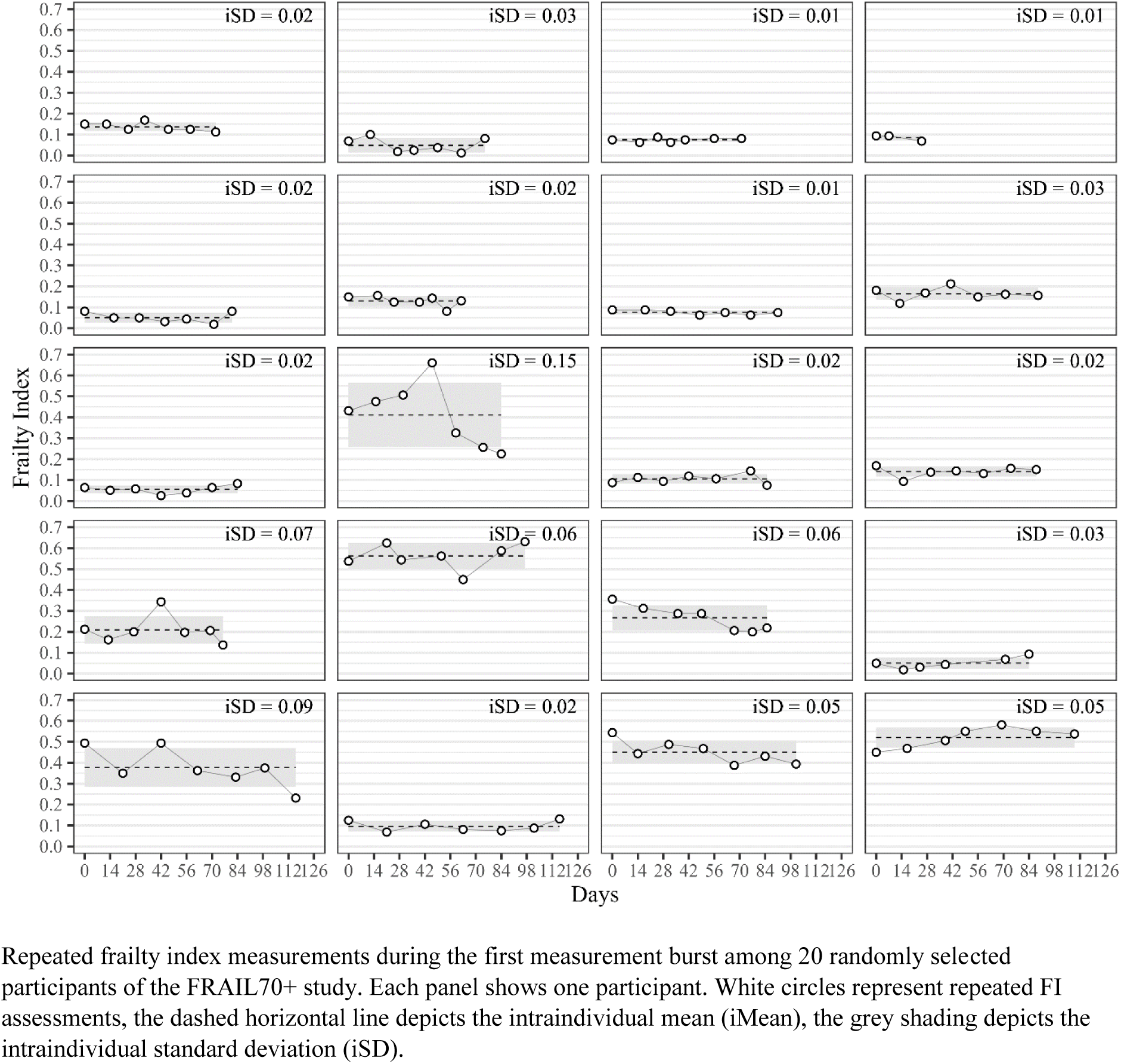
Repeated frailty index assessments among 20 randomly selected older adults.

**Figure 2:**
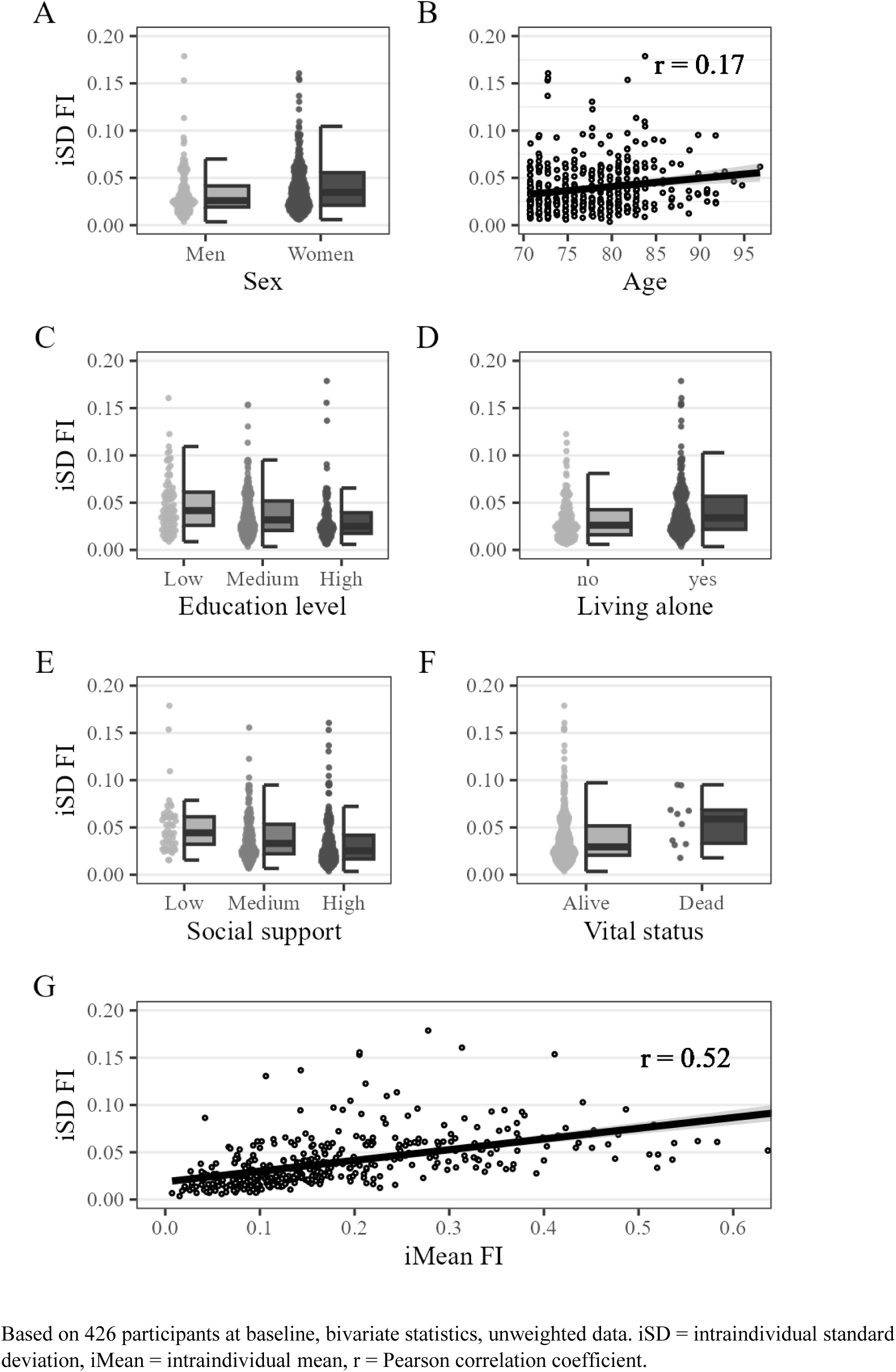
Bivariate characteristics of intraindividual variability (iSD) in the frailty index (FI)

**Figure 3:**
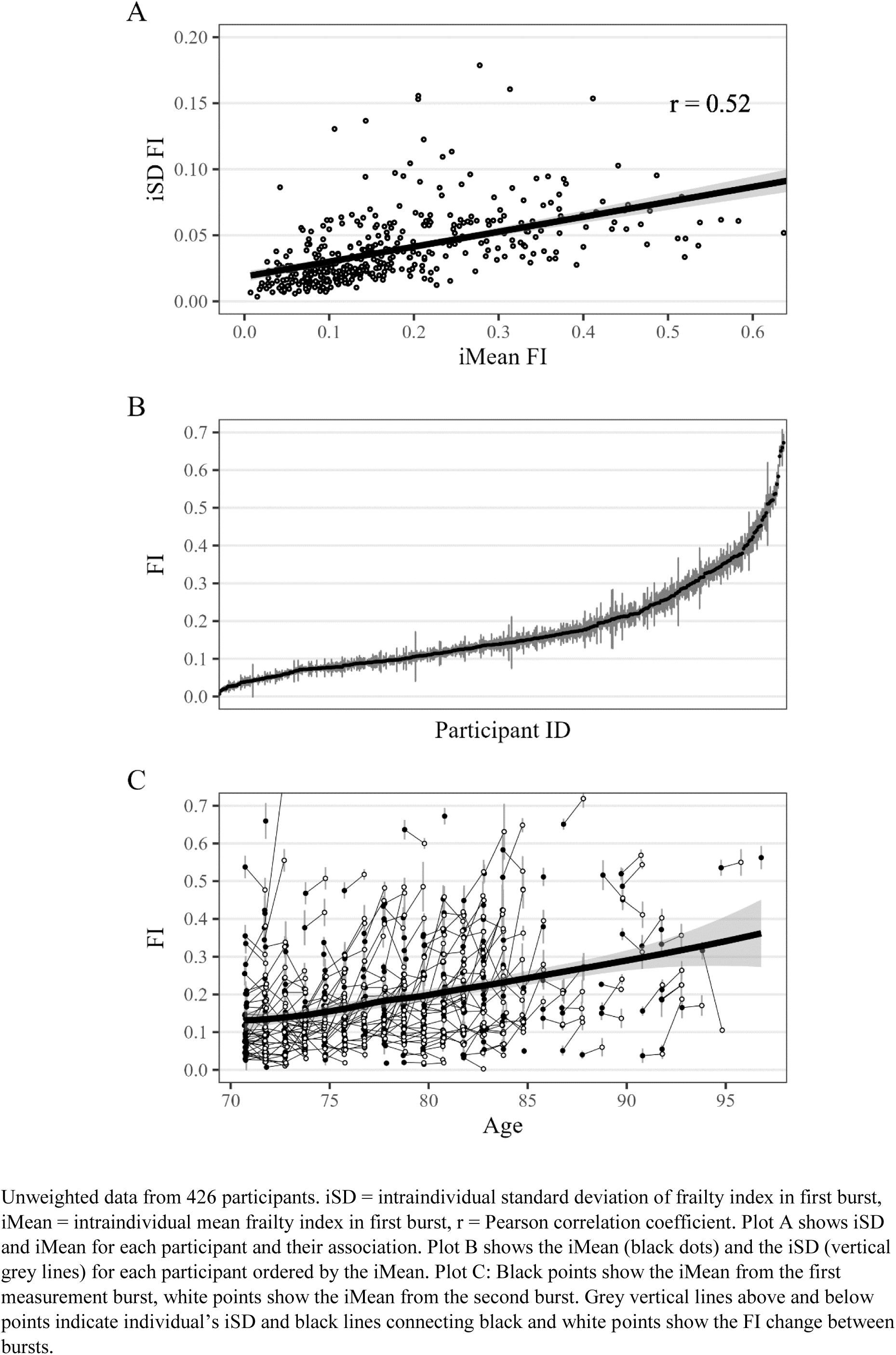
Relationship between average frailty index (FI) level and short-term FI fluctuations.

Next step, we assessed where short-term FI fluctuations originated from by calculating the iIQR for each health deficit, which depicts the middle 50% of within-burst intraindividual variability. Supplementary Figure 4 shows that person- and deficit-level iIQRs were sparsely distributed with a strong zero-inflation, that is, most older adults had no change within bursts per each deficit. Supplementary Table 2 shows for each health deficit, the proportion of participants who had none, some (iIQR≤0.5), or high (iIQR>0.5) within-person fluctuations. Expectedly, none or very limited change shows among self-reported chronic diseases and conditions. Smaller within-burst changes were relatively common for deficits with more categories such as self-rated health, tiredness, and sleeping or hearing problems, but a few dichotomous deficits stood out as being particularly unstable: physical inactivity, dizziness, and difficulties with dressing, shopping, walking 100 meters, climbing stairs, or carrying 5kg as well as both cognitive deficits (attention, memory). The three health deficits depicting negative affect (depressed, sad, lonely) also showed some fluctuation. Finally, among the subsample with repeated in-person interviews (Supplementary Figure 5), we found that deficits based on physical performance tests also showed considerable biweekly fluctuations, which fits with the heightened fluctuations in self-reported mobility-related limitations.

Next step, we compared individual’s FI fluctuations across different chronic diseases to assess whether FI fluctuations reflect disease-specific fluctuating symptomatology. Supplementary Figure 6 shows that for older adults with heart disease, lung disease, diabetes, osteoarthritis, and dementia, the iSD was 0.010-0.015 (+20-40%) higher compared to those without. No differences were found for the remaining chronic diseases (hypotension, stroke, cancer, renal disease).

Results from the unconditional location-scale mixed regression model (M0) indicated, that from the total FI variance, 77.3% was attributable to stable (over 1.5 years) between-person FI differences, 9.8% to across burst/within-person long-term FI changes, and the remaining 12.9% due to within-burst/within-person short-term FI variability. In other words, short-term FI fluctuations accounted for bit more variance than 1-year FI changes. Comparing the unconditional location-only (M1) with the unconditional location-scale model (M2, Supplementary Table 3) shows a clear improvement in model fit, i.e., incorporating short-term FI fluctuations provides a better representation of the data. Results from model 2 (Supplementary Table 3) reiterate the close interrelationship between an older person’s average level of frailty as well as their one-year change with short-term fluctuations: the FI level correlated strongly (r=0.79) and FI change moderately (r=0.37) with FI fluctuations. In other words, older adults who were frailer overall and whose health deteriorated more during follow-up also reported more unstable health.

Results from the final location-scale model (M4, Table 1) show that FI fluctuations increased with baseline age (+3% per year), were higher among women (+15%) than men, higher for those with low compared to those with high education (+39%), higher for those with low compared to high social support (+47%), as well as those who died between bursts (+44%) compared to survivors. Older adults who experienced a negative health event during a burst, that is, those who took to bed due to a health problem (+50%) and those who had an overnight hospital stay (+50%) had more FI fluctuations. Experiencing a fall, on the other hand, was not consistently associated with higher FI fluctuations over the preceding or following weeks in the full model.

**Table 1:** Results from final location-scale mixed regression model.

|  | Frailty level | Frailty fluctuations |
| --- | --- | --- |
| | $\mu$ (95%-CI) | $\sigma$ (95%-CI) |
| FIXED EFFECTS |  |  |
| Intercept | 0.27 (0.23, 0.31) | 0.04 (0.04, 0.06) |
| Wave (within-burst) | -0.00 (-0.00, 0.00) | - |
| FI change (burst 2 vs. burst 1) | 0.01 (0.01, 0.02) | 0.98 (0.91, 1.05) |
| Age (years) | 0.01 (0.01, 0.01) | 1.03 (1.02, 1.04) |
| Women (vs. men) | 0.02 (-0.01, 0.04) | 1.15 (1.02, 1.29) |
| Medium education (vs. low) | -0.04 (-0.07, -0.01) | 0.80 (0.69, 0.92) |
| High education (vs. low) | -0.07 (-0.10, -0.04) | 0.72 (0.61, 0.84) |
| Living alone (vs. cohabiting) | 0.02 (-0.00, 0.04) | 1.12 (1.00, 1.26) |
| Medium social support (vs. low) | -0.08 (-0.12, -0.05) | 0.80 (0.69, 0.92) |
| High social support (vs. low) | -0.11 (-0.14, -0.08) | 0.68 (0.57, 0.80) |
| Dead (vs. alive in burst 2) | 0.17 (0.10, 0.24) | 1.44 (0.94, 2.13) |
| Bedrest (vs. none) | 0.04 (0.03, 0.04) | 1.50 (1.36, 1.65) |
| Fall injury (vs. none) | 0.01 (0.00, 0.01) | 1.14 (0.97, 1.33) |
| Hospital stay (vs. none) | 0.03 (0.02, 0.06) | 1.50 (1.12, 1.97) |
| RANDOM EFFECTS |  |  |
| Individual-level intercept (SD) | 0.10 (0.09, 0.11) | 0.02 (0.02, 0.03) |
| Individual-level intercepts (COR) | 0.62 (0.52, 0.72) |  |
| Burst-level intercept (SD) | 0.03 (0.02, 0.03) | 0.36 (0.32, 0.41) |
| Burst-level wave (SD) | 0.00 (0.00, 0.00) | - |
| Burst-level intercepts (COR) | 0.26 (0.07, 0.44) |  |
| Burst-level intercept*wave (COR) | -0.35 (-0.56, 0.09) | 0.21 (-0.05, 0.47) |
| MODEL FIT |  |  |
| LOO | -19,432 |  |
| R<sup>2</sup> (fixed effects only) | 0.293 |  |

Results from final location-scale mixed regression model (model 4) based on unweighted data from 426 participants and 5,122 observations, adjusted for interview mode. $\mu$ refers to the coefficients of the location sub-model, $\sigma$ to the (exponentiated) coefficients of the scale sub-model. 95%-CI = 95% credible intervals, SD = standard deviation, R<sup>2</sup> = Bayesian R-squared based on fixed effects, LOO=leave-one-out cross-validation.

## DISCUSSION

In this study, we differentiated empirically between (1) stable FI differences between older adults, (2) durable long-term within-person FI changes, and (3) reversible short-term within-person FI fluctuations. We found that short-term FI fluctuations were neither negligibly small nor random. Short-term FI fluctuations increased with chronological age, the degree of frailty, frailty change, were higher among those who died during follow-up, were related with some chronic diseases, and partially reflected discrete health events. In conclusion, we consider intraindividual variability in the FI to not only contain unintelligible and random noise, but also systematic information about the aging process at the person level or at least the measurement thereof.

Based on repeated biweekly assessments in two measurement bursts one year apart, we found fluctuations in the clinical FI to amount to about one third of the between-person differences at baseline, which is substantial but lower than what previous studies^48,59^ on intraindividual variability in cognitive and physical functioning have reported. This is not surprising as previous studies have focussed (only) on more dynamic (cognitive) functioning, while the standard clinical FI includes a number of chronic diseases which were as expected mostly stable across weeks and months. On an absolute scale, FI fluctuations in our study amounted to 0.04, which is very close to a previous estimate^32^ of FI fluctuations. This confirmation is reassuring, particularly as the previous estimate was based on more extensive cross-national but at the same time sparser (less intensive) biannual data. In the current study, short-term FI fluctuations were fuelled particularly by fluctuations in cognitive health deficits, somatic symptoms, and limitations in instrumental and mobility-related activities of daily living. Smaller fluctuations were prevalent in a number of health deficits including those depicting sensory impairment, pain, negative affect, sleep problems, and fatigue. These findings are broadly compatible with previous research reporting substantial fluctuations in sleep time^46^, pain^45^, cognition^48^ and disability^47^ over days, weeks, and months. Short-term changes in disability have been reported before^60–62^, although we found here that this applied particularly to limitations in the more challenging instrumental and mobility-related rather than basic activities of daily living.

It could be argued that the weekly short-term fluctuations we found are mainly due to inherent limitations of self-reports, which include coarse categories, biased reporting^63,64^ or problems to reliably detect change^65^. However, the overall FI is a fine-grained and generally reliable^33,34^ tool to differentiate between older adults, and in our study, we found short-term fluctuations not only in self-reported health problems, but also in those based on more objective physical performance tests, which is in line with two small-scale studies^59,66^ that also reported weekly fluctuations in mobility-related physical performance tests.

Existing small-scale studies also indicate the close relationship between cognitive and physical performance: older adults with central nervous system dysfunction and more fluctuations in cognition also tend to have more unstable physical functioning. Strauss et al.^66^ concluded that “*measures of cognitive as well as physical* [intraindividual] *variability are important behavioural markers of neurological integrity*”. In the same study, fluctuations in negative affect, were associated on the other hand with other more transient processes such as pain variability. This is compatible with our study results, where we also found more fluctuations among older adults with osteoarthritis and dementia, and some variability in negative affect, which don’t seem to be the main source of fluctuations in the overall FI. Given the nature of the FI as a summary index, it is unclear though how fluctuations in the constituting health deficits – and the underlying phenomena – align and interact with each other over time, which should be addressed more systematically in future research, using for example a network analysis approach^67,68^.

On the level of the overall FI, the association of fluctuations with older adults’ chronological age, frailty level as well as frailty change and mortality risk implies that short-term FI fluctuations are related to the aging process, similar to fluctuations in cognitive and physical functioning^59^. As older adults age, they do not only accumulate more and more physical and cognitive health problems^12^ that then result in lower average levels of functioning in the physical and cognitive domain^69^ – particularly towards the end of life^70^ – but their level of functioning also becomes more unstable over time, with some deficits blinking in and out of the set of accumulated health problems. Importantly, we could show that short-term fluctuations were not only higher among older adults who were older and frailer but also among those whose average level of frailty increased *during* the study.

Our results are compatible with three mechanisms linking reversible fluctuations in the FI and aging: First, both mild and severe cognitive impairment have been associated with variability in cognitive functioning^35,59,71^, the latter which has also been linked to variability in physical functioning^66^. Second, short-term fluctuations may also arise due to variability in somatic symptoms like pain and fatigue, which might partly be disease-specific and which may give rise to affect variability^66^. Third, with age, the probability for discrete negative health events like injuries, severe cases of infection and hospital stays increases. These, together with subsequent recoveries^38,40^ can give rise to fluctuations in the overall FI over weeks and months.

Our results for older adults imply that as they age, their level of functioning becomes more unstable and hence less predictable, which might negatively affect short- and medium-term plannability of activities. It would be interesting to assess, whether the reported and estimated instability in health status has a negative real-world impact on older adults’ quality of life above and beyond the average health status, and if so, whether instability in functioning and somatic symptoms can be reduced by interventions. An implication of our results for both researchers and clinicians is that a one-time FI assessment should be seen as just one data point in a long string of potential FI measurements, and that these become more unstable as older adults become frailer. Hence, the results of one-time assessments should not be over-interpreted, particularly concerning whether someone is ‘robust’ or ‘frail’. Our results imply that broad classification based on a cut-off applied to a one-time measurement is associated with a risk of miss-classification, as older adult’s FI may fluctuate across thresholds. Therefore, we suggest using finely-grained continuous data provided by the FI, i.e., the degree of frailty^13^, rather than insisting on coarse categorization, which is something that also extends to the FI-constituting health deficits.^72,73^ Our results also have implications with regard to the selection of health deficits for clinical FIs: including more comorbidities – similar to FIs based on electronic claims or patient data^74,75^ – will reduce short-term fluctuations in clinical FIs^76^, but this needs to be balanced with the added value^34,68^ of health deficits reflecting overall health and those covering cognitive and physical functioning, so that the FI does not merely reflect multimorbidity^55^.

Our results also indicate that previously reported clinically meaningful changes for the FI^50,51^ are easily within range of reversible short-term FI fluctuations over weeks, particularly if this person has already a high level of frailty. Hence, while the FI reliably differentiates *between* older adults, its capacity for precise longitudinal monitoring, that is, *within* older individuals is more questionable.^34^ As ‘normal’ fluctuations^48^ increase with age and the degree of frailty, monitoring durable FI changes becomes more difficult. Hence, what constitutes a meaningful change in the FI should be calibrated based on where adults’ are located on the robustness-frailty continuum. That is, the same absolute amount of change in the FI is more salient in a robust compared to a frail older person whose fluctuations are larger. Of course, this is not to say that frail (vulnerable) older adults should not be monitored, but only that extrapolating a durable health deterioration (or improvement) from an apparent change in the clinical FI is more difficult. One option for frailty monitoring that circumvents being led astray by short-term fluctuations is to rely on estimated FI levels and trajectories derived from analytic procedures that smooth over intraindividual variability^77,78^ rather than to use the more unstable raw observed FI values.

To our knowledge, this is the first paper systematically assessing intraindividual variability or short-term fluctuations in the FI. Next to the strengths of its nationwide sample, the measurement burst design with many repeated biweekly assessments per person and the differentiation between stable between-person differences, durable within-person change and reversible within-person fluctuations, there are also several limitations. First, health deficits for the FI were generally self-reported, hence some of the biweekly variability likely comes from misunderstanding and misremembering as well as context effects during the interviews, and the coarse nature of the instruments. However, previous studies suggest that FIs based on self-reports are comparable to those based on test-based health deficits^79^. Similarly, variability in cognitive and physical performance tests as well as the size and systematic properties imply that there is more to FI fluctuations than just random measurement error or noise. Second, this study examined community-dwelling older adults only. Given the association with the frailty level, we would expect that short-term FI fluctuations are likely substantially higher among hospitalised and institutionalised older adults. Third, very few participants died during follow-up, hence it was not possible to test whether FI fluctuations predict negative outcomes beyond the average FI level. However, given the high correlation with the average FI and the additional effort required to assess short-term fluctuations, this seems both unlikely and uneconomic. Rather than a causal or predictive factor, our results imply that fluctuations in the FI, that is, ups and downs in older adults’ overall health status across weeks and months, are a concomitant phenomenon of the aging process, of which researchers and clinicians should be aware. In contrast to the unclear prognostic utility of short-term fluctuations, we think that repeated FI assessments over longer periods (annually or biannually) depicting long-term changes or trajectories can provide a benefit over one-time assessments.^77,80,81^

## CONCLUSION

Short-term FI fluctuations were neither small nor random. Instead, as older adults become frailer, their measured health also becomes more unstable, hence short-term fluctuations in older adults’ health status due to fluctuations in cognitive and physical functioning, somatic symptoms, and health events can be seen as a concomitant phenomenon of aging. Researchers and clinicians should be aware of the existence of short-term FI fluctuations and their consequences for frailty monitoring.

## Author contributions

E.S. is the corresponding author, he planned the study, performed all statistical analysis, and wrote the article. A.S. reviewed the R-code and critically reviewed the manuscript. E.O.H, O.T, and K.R. critically reviewed the manuscript.

## Data availability

Data of the FRAIL70+ study are freely available for researchers via the Austrian Social Science Data Archive (AUSSDA: https://doi.org/10.11587/DJNOHX) and the R-code to reproduce all analyses and results are also available online (https://osf.io/p6xwh/).

## Supplementary Material

Supplementary Figure 1: Assessment schedule FRAIL70+ study

Supplementary Table 1: Health deficits of frailty index

Supplementary Methods 1: Sampling procedure

Supplementary Figure 2: Frailty index characteristics (at baseline)

Supplementary Methods 2: Description of performance tests of physical function

Supplementary Methods 3: Statistical analysis

Supplementary Figure 3: Descriptive statistics for frailty index (FI) in both measurement bursts by sex

Supplementary Figure 4: Distribution of intraindividual variability (iSD) values by health deficits of frailty index (first burst)

Supplementary Table 2: Short-term fluctuations at health deficit level: proportion of intraindividual interquartile range (iIQR) categories

Supplementary Figure 5: Distribution of intraindividual variability (iSD) values by health deficits of frailty index (first burst) among subgroup of participants including physical performance tests

Supplementary Figure 6: Short-term frailty index fluctuations (iSD) by chronic disease status

Supplementary Table 3: Results from mixed regression models

