## Supplementary Material for "Short-term frailty index fluctuations in older adults: Noise or signal?"

Supplementary Figure 1: Assessment schedule FRAIL70+ study

Supplementary Table 1: Health deficits of frailty index

Supplementary Methods 1: Sampling procedure

Supplementary Figure 2: Frailty index characteristics (at baseline)

Supplementary Methods 2: Description of performance tests of physical function

Supplementary Methods 3: Statistical analysis

Supplementary Figure 3: Descriptive statistics for frailty index (FI) in both measurement bursts by sex

Supplementary Figure 4: Distribution of intraindividual variability (iSD) values by health deficits of frailty index (first burst)

Supplementary Table 2: Short-term fluctuations at health deficit level: proportion of intraindividual interquartile range (iIQR) categories

Supplementary Figure 5: Distribution of intraindividual variability (iSD) values by health deficits of frailty index (first burst) among subgroup of participants including physical performance tests

Supplementary Figure 6: Short-term frailty index fluctuations (iSD) by chronic disease status

Supplementary Table 3: Results from mixed regression models

Supplementary Figure 1: Assessment schedule FRAIL70+ study

Burst 1

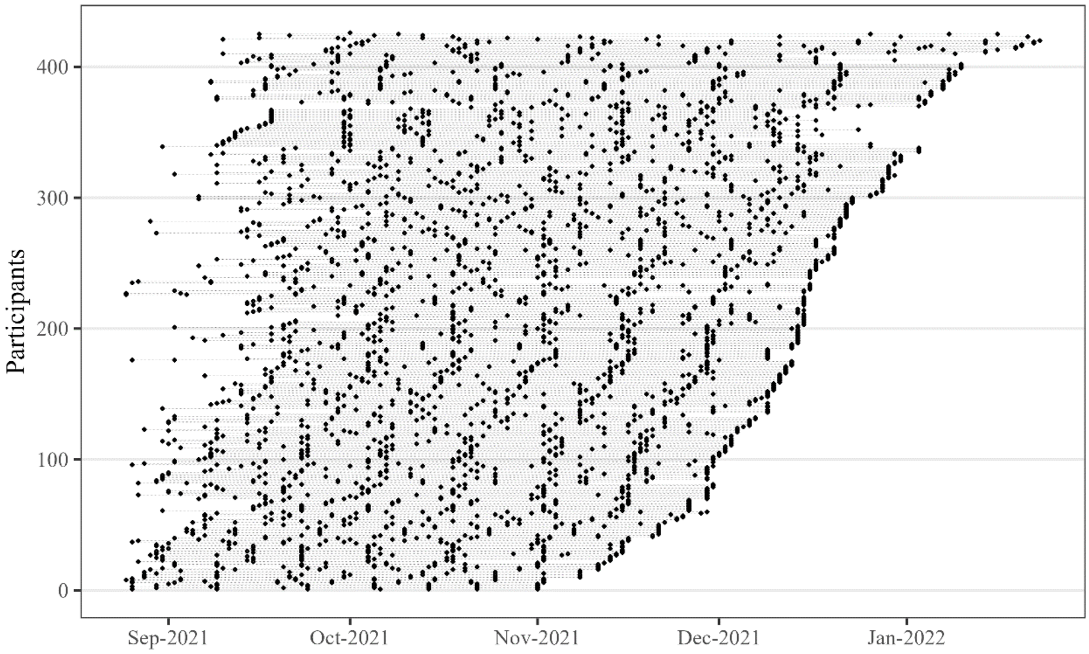

Burst 2

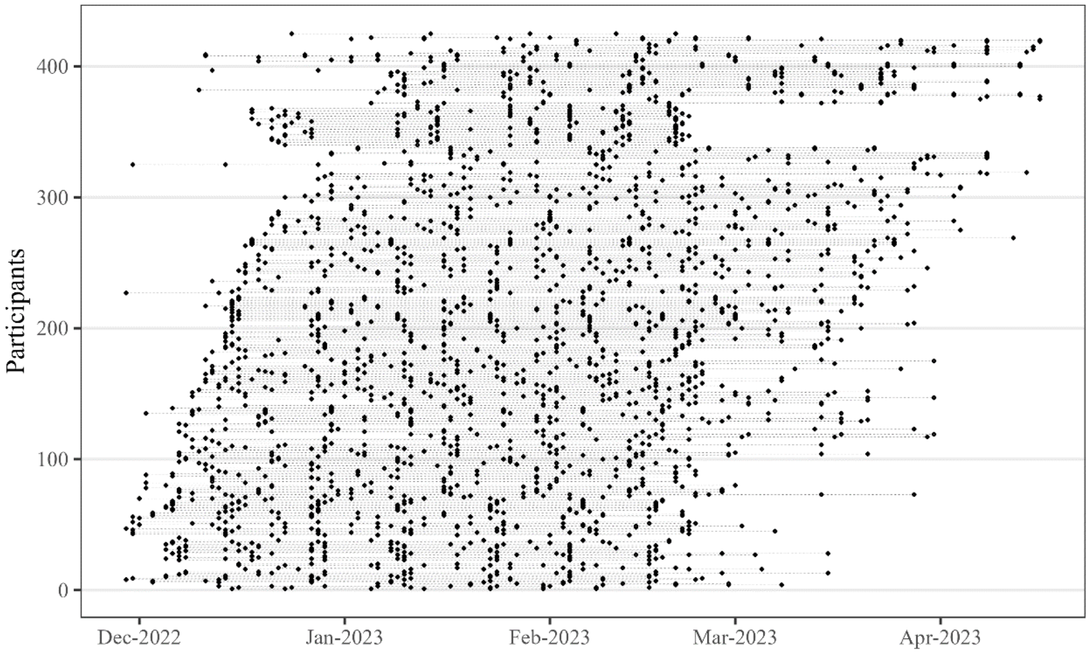

Black dots refer to assessments, dashed grey lines represent individuals.

Supplementary Table 1

| Health deficit | Coding | Prevalence at baseline in % | Missing data at baseline in % |
|---|---|---|---|
| Self-rated health | excellent = 0, very good = 0.25, good = 0.50, moderate = 0.75, poor = 1 | 0 = 6.3<br>0.25 = 18.5<br>0.50 = 35.9<br>0.75 = 28.4<br>1 = 10.8 | - |
| Dizziness | no = 0, yes = 1 | 1 = 20.7 | - |
| Pain | pain rating from 0-10. 0 = 0, ≥ 1 & ≤ 3 = 0.5, ≥ 4 = 1 | 0 = 26.1<br>0.5 = 35.2<br>1 = 38.7 | - |
| Tiredness | never = 0,<br>sometimes = 0.5,<br>always/often = 1 | 0 = 43.0<br>0.5 = 42.0<br>1 = 15.0 | - |
| Vision | excellent = 0, very good = 0.25, good = 0.50, moderate = 0.75, poor = 1 | 0 = 10.4<br>0.25 = 38.0<br>0.50 = 35.1<br>0.75 = 13.4<br>1 = 3.1 | 0.5 |
| Hearing | excellent = 0, very good = 0.25, good = 0.50, moderate = 0.75, poor = 1 | 0 = 12.7<br>0.25 = 36.2<br>0.50 = 30.8<br>0.75 = 16.9<br>1 = 3.3 | 0.2 |
| Attention | 10 words immediate recall test.<br>≥ 5 = 0, < 5 = 1 | 1 = 20.0 | - |
| Memory | 10 word delayed recall test.<br>≥ 4 = 0, < 4 = 1 | 1 = 32.6 | - |
| Physical inactivity | moderate physical activity: “Every day/almost every day” and “multiple times a week” = 0, “once per week” & “less often” = 1 | 1 = 21.4 | - |
| Doctor told you had: Heart problem (myocardial infarction, coronary thrombosis, other problem including congestive heart failure) | no = 0, yes = 1 | 1 = 15.3 | - |
| Doctor told you had: High blood pressure or hypertension | no = 0, yes = 1 | 1 = 48.6 | - |
| Doctor told you had: Stroke or cerebral vascular disease | no = 0, yes = 1 | 1 = 4.7 | - |
| Doctor told you had: Diabetes or high blood sugar | no = 0, yes = 1 | 1 = 19.5 | - |
| Doctor told you had: Chronic lung disease such as chronic bronchitis or emphysema | no = 0, yes = 1 | 1 = 9.9 | - |
| Doctor told you had: Cancer or malignant tumour, including leukaemia or lymphoma | no = 0, yes = 1 | 1 = 5.6 | - |
| Doctor told you had: Arthritis, including osteoarthritis, or rheumatism | no = 0, yes = 1 | 1 = 27.0 | - |
| Doctor told you had: Chronic renal disease | no = 0, yes = 1 | 1 = 2.8 | - |
| Doctor told you had: Alzheimer's disease, dementia or any other serious memory impairment | no = 0, yes = 1 | 1 = 3.1 | - |
| Difficulty dressing | no = 0, yes = 1 | 1 = 12.2 | - |
| Difficulty walking across room | no = 0, yes = 1 | 1 = 8.0 | 0.5 |
| Difficulty bathing/showering | no = 0, yes = 1 | 1 = 9.9 | - |
| Difficulty eating | no = 0, yes = 1 | 1 = 3.8 | - |
| Difficulty going in/out of bed | no = 0, yes = 1 | 1 = 7.5 | - |
| Difficulty using toilet | no = 0, yes = 1 | 1 = 3.5 | - |
| Difficulty preparing a warm meal | no = 0, yes = 1 | 1 = 5.4 | - |
| Difficulty shopping groceries | no = 0, yes = 1 | 1 = 11.8 | 0.5 |
| Difficulty using telephone | no = 0, yes = 1 | 1 = 1.6 | 0.7 |
| Difficulty taking medicine | no = 0, yes = 1 | 1 = 1.9 | 1.6 |
| Difficulty walking 100 meters | no = 0, yes = 1 | 1 = 12.5 | 0.7 |

| | | | |
|---|---|---|---|
| Difficulty taking one flight of stairs | no = 0, yes = 1 | 1 = 23.6 | 0.7 |
| Difficulty reaching or extending your arms above shoulder level | no = 0, yes = 1 | 1 = 14.8 | - |
| Difficulty lifting or carrying weights over 10 pounds/5 kilos, like a heavy bag of groceries | no = 0, yes = 1 | 1 = 27.8 | 0.5 |
| Lonely | never/rarely = 0<br>sometimes = 0.5<br>often/always = 1 | 0 = 76.8<br>0.5 = 17.6<br>1 = 5.6 | - |
| Difficulty concentrating | never/rarely = 0<br>sometimes = 0.5<br>often/always = 1 | 0 = 71.4<br>0.5 = 25.8<br>1 = 2.8 | - |
| Depressed | never/rarely = 0<br>sometimes = 0.5<br>often/always = 1 | 0 = 69.0<br>0.5 = 26.1<br>1 = 4.9 | - |
| Everything takes effort | never/rarely = 0<br>sometimes = 0.5<br>often/always = 1 | 0 = 66.9<br>0.5 = 23.9<br>1 = 9.2 | - |
| Sleep problems | never/rarely = 0<br>sometimes = 0.5<br>often/always = 1 | 0 = 52.7<br>0.5 = 35.3<br>1 = 12.0 | 0.2 |
| Could not get going | never/rarely = 0<br>sometimes = 0.5<br>often/always = 1 | 0 = 65.7<br>0.5 = 27.7<br>1 = 6.6 | - |
| Sad | never/rarely = 0<br>sometimes = 0.5<br>often/always = 1 | 0 = 72.3<br>0.5 = 23.0<br>1 = 4.7 | - |
| Poor appetite | never/rarely = 0<br>sometimes = 0.5<br>often/always = 1 | 0 = 89.0<br>0.5 = 8.0<br>1 = 3.1 | - |

### Supplementary Methods 1: Sampling procedure

The contracted survey agency IFES (Institut für empirische Sozialforschung, Vienna) contacted 971 community-dwelling older adults aged 70 year and above based on previous participation in population-representative studies of the agency, which includes, for example, the Austrian participants of the Survey of Health, Ageing and Retirement in Europe (SHARE). Out of the 971 selected individuals, 426 agreed to participate, which amounts to a response rate of 44%. Before participation, interviewers described the topic, length, and required information of the study, ensured anonymity of all personal data, and obtained written consent for participation.

When selected older adults were successfully contacted but did not want to participate in the full study, they were asked to provide basic demographic and health-related information, to provide a better understanding of selection into the sample. In comparison to participants, those who could or would not participate were more likely men (42.8% vs. 35.4%; $\chi^2=5.3$, df=1, p=0.021), had only minimum compulsory schooling (28.1% vs. 19.2%; $\chi^2=20.3$, df=2, p<0.001), and had poorer self-reported health (moderate = 37.5% vs. 28.4%, poor = 11.1% vs. 10.8%; $\chi^2=23.9$, df=4, p<0.001), but were of similar age (mean=77.7 vs. 77.3 years; F-statistic=0.76, df=1, p=0.383). Compared to the general older population 70+ years or over in Austria based on information from Statistics Austria, women (65% instead of 58%) and those who finished upper secondary or tertiary education (27% instead of 14%) were overrepresented in the FRAIL70+ study. Also, the sample was on average half a year younger (mean age=77.7 years) than the overall population (78.3 years).

Supplementary Figure 2: Frailty index characteristics (at baseline)

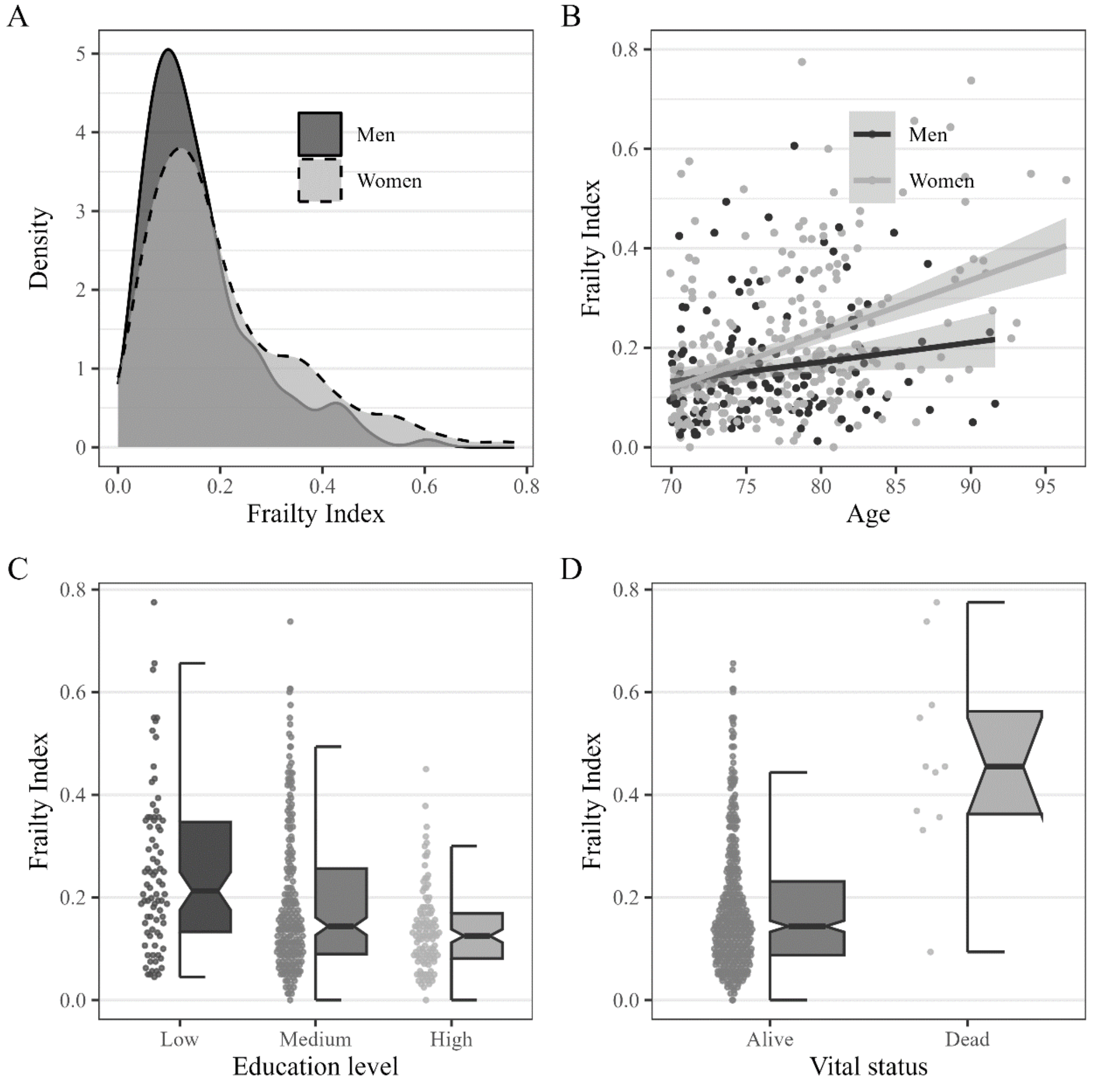

Supplementary Methods 2: Description of health deficits in physical performance tests

Slow walking speed was measured by belonging to the lowest 20% in normal gait speed in seconds over 2.5 meter based on the lower value over two trials. Slow chair rise test scores were based on the time in seconds for five repeated chair rises (>14 seconds for those aged ≤ 79 years and > 16 seconds for those aged 80+ years). Muscle weakness was based on previously specified sex- and BMI-specific grip strength values (men: GS ≤ 29 & BMI ≤ 24 = 1, GS ≤ 30 & BMI >24 & BMI ≤ 28 = 1, GS ≤ 32 & BMI > 28 = 1; women: GS ≤ 17 & BMI ≤ 23 = 1, GS ≤ 17.3 & BMI >23 & BMI ≤ 26 = 1, GS ≤ 18 & BMI > 26 & BMI ≤ 29 = 1, GS ≤ 21 & BMI > 29 = 1) measured as the maximum reading (in kg) over four trials (two per each hand) with a handheld dynamometer (Smedley S Dynamometer, TTM, Tokyo, 100kg). Participants who could not perform the walking speed, chair rise, or grip strength test were considered to fulfil the respective deficit criterion.

Supplementary Methods 3: Statistical analysis

For this analysis, a three-level mixed regression approach was used to account for the nesting of repeated FI assessments (level 1) within bursts (level 2) within individuals (level 3). The first model was a location-only unconditional means model which we used to calculate variance components, that is, to ascertain how much variance of the FI is attributable to stable (over 1.5 years) between-person differences (FI level), to within-person/between-burst change (one-year FI change), and to within-person/within-burst biweekly short-term FI fluctuations. This model (M0) can be written as:

$$\begin{aligned} FI_{tij} &= \pi_{0ij} + \epsilon_{tij} \\ \pi_{0ij} &= \beta_{00j} + r_{0ij} \\ \beta_{00j} &= \gamma_{000} + u_{00j} \\ r_{0ij} &\sim N(0, \tau_{00}) \\ u_{00j} &\sim N(0, \tau_{00}) \\ \epsilon_{tij} &\sim N(0, \sigma^2) \end{aligned}$$

The first line depicts level 1, where $FI_{tij}$ is the observed Frailty Index score at time-point $t$ in burst $i$ for person $j$. $\pi 0_{ij}$ refers to the average FI per burst and person. The level-2 equation states that $\pi_{0ij}$ is equal to the average FI per burst ($\beta_{00j}$) plus an individual-specific deviation ($r_{0ij}$) therefrom. The level 3 equation in the third line states that individual's FI is a combination of the overall FI ($\gamma_{000}$) and the individual's deviation ($u_{00j}$) therefrom. The deviations from intercepts across both bursts ($r_{0ij}$) and individuals ($u_{00j}$) are random effects which are not directly estimated but assumed to follow a multivariate normal distribution with a mean of zero and an estimated variance matrix ($\tau_{00}$). Note that there are no fixed effects (that is, fixed across bursts or individuals) besides the overall intercept in this unconditional means location-only model. Finally, there is the within-burst and within-person variance ($\varepsilon_{tij}$) – that is, the variance of short-term FI fluctuations – which is again not directly estimated but assumed to stem from a normal distribution with a mean of zero and an estimated variance. Based on the two estimated random effect variances and the residual variance, we calculated the intraclass correlation coefficient (ICC) for each level as the proportion of the sum of all three variance components.

In the second unconditional model, we added the time variables within-burst assessment and burst to the second-level equation:

$$\begin{aligned} FI_{tij} &= \pi_{0ij} + \epsilon_{tij} \\ \pi_{0ij} &= \beta_{00j} + \beta_1 Burst_{ij} + \beta_2 Assessment_{ij} + \\ &\beta_3 InterviewMode_{ij} + r_{0ij} + r_{1ij} \\ \beta_{00j} &= \gamma_{000} + u_{00j} \\ r_{0ij} &\sim N(0, \tau_{00}) \\ u_{00j} &\sim N(0, \tau_{00}) \\ \epsilon_{tij} &\sim N(0, \sigma^2) \end{aligned}$$

Burst ($\beta_1$) denotes the 1-year FI change, and assessment ($\beta_2$) was added to ensure the de-trending of repeated FI assessments, that is, to meet the key assumption that within-person FI assessments are independent from each other (and from time). For this, we also added assessment as random slope effect ($r_{1ij}$) to allow for individual-specific trends. The main purpose of M1 is to serve as the baseline model for comparison with the next model, where we added the scale-component, i.e., when we allowed the within-individual within-burst residual error to vary across individuals and to be explicitly modelled. This unconditional location-scale growth curve model (M2) is formulated as follows:

$$FI_{tij} = \pi_{0ij} + \epsilon_{tij}$$
$$\pi_{0ij} = \beta_{00j} + \beta_1 Burst_{ij} + \beta_2 Assessment_{ij} +$$
$$\beta_3 InterviewMode_{ij} + r_{0ij} + r_{1ij}$$
$$\beta_{00j} = \gamma_{000} + u_{00j}$$
$$r_{0ij} \sim N(0, \tau_{00})$$
$$u_{00j} \sim N(0, \tau_{00})$$
$$\sigma_j^2 = exp[w_{00j} + w_1 Burst_{ij} + w_2 InterviewMode_{ij} + r_{0ij} + u_{00j}]$$

The key modification is in line 6, which allows the individual-specific residual variance to become a second outcome. Here, we specified the average residual variance ($w_{00j}$), that is, short-term FI fluctuation, and the individuals' deviation therefrom. Within-person/within-burst residual variance is estimated with a log-linear model to ensure that the estimated variance is non-negative. Exponentiating this term later on leads to a multiplicative interpretation of the predictors of FI fluctuations (i.e., in terms of % differences). Importantly, the random effect are allowed to correlate both within and across location and scale sub-models, which provides insights into how FI level and FI change relate to FI fluctuations.

In the next model (M3), we included time-constant predictor variables for both FI level and FI fluctuations:

$$FI_{tij} = \pi_{0ij} + \epsilon_{tij}$$
$$\pi_{0ij} = \beta_{00j} + \beta_1 Burst_{ij} + \beta_2 Assessment_{ij} +$$
$$\beta_3 InterviewMode_{ij} + r_{0ij} + r_{1ij}$$
$$\beta_{00j} = \gamma_{000} + \gamma_{001} Age_j + \gamma_{002} Female_j + \gamma_{003} MediumEDU_j +$$
$$\gamma_{004} HighEDU_j + \gamma_{005} Alone_j + \gamma_{006} MediumSUPP_j + \gamma_{007} HighSUPP_j +$$
$$\gamma_{008} Died_j + u_{00j}$$
$$r_{0ij} \sim N(0, \tau_{00})$$
$$u_{00j} \sim N(0, \tau_{00})$$
$$\sigma_j^2 = exp[w_{00j} + w_1 Burst_{ij} + w_2 InterviewMode_{ij} + w_{001} Age_j +$$
$$w_{002} Female_j + w_{003} MediumEDU_j + w_{004} HighEDU_j + w_{005} Alone_j +$$
$$w_{006} MediumSUPP_j + w_{007} HighSUPP_j + w_{008} Died_j + r_{0ij} + u_{00j}]$$

In the final model (M4), we added three-time-varying predictors for both the location and the scale sub-model:

$$FI_{tij} = \pi_{0ij} + \epsilon_{tij}$$
$$\pi_{0ij} = \beta_{00j} + \beta_1 Burst_{ij} + \beta_2 Assessment_{ij} +$$
$$\beta_3 InterviewMode_{ij} + r_{0ij} + r_{1ij}$$
$$\beta_{00j} = \gamma_{000} + \gamma_{001} Age_j + \gamma_{002} Female_j + \gamma_{003} MediumEDU_j +$$
$$\gamma_{004} HighEDU_j + \gamma_{005} Alone_j + \gamma_{006} MediumSUPP_j + \gamma_{007} HighSUPP_j +$$
$$\gamma_{008} Died_j + \gamma_{009} Bedrest_{tij} + \gamma_{010} Fall_{tij} + \gamma_{011} Hospital_{tij} + u_{00j}$$
$$r_{0ij} \sim N(0, \tau_{00})$$
$$u_{00j} \sim N(0, \tau_{00})$$
$$\sigma_j^2 = exp[w_{00j} + w_1 Burst_{ij} + w_2 InterviewMode_{ij} + w_{001} Age_j +$$
$$w_{002} Female_j + w_{003} MediumEDU_j + w_{004} HighEDU_j + w_{005} Alone_j +$$
$$w_{006} MediumSUPP_j + w_{007} HighSUPP_j + w_{008} Died_j + w_{009} Bedrest_{tij} +$$
$$w_{010} Fall_{tij} + w_{011} Hospital_{tij} + r_{0ij} + u_{00j}]$$

For model comparison, we used leave-one-out cross-validation (LOO) where lower values imply better model fit. For absolute model fit, we calculated the Bayesian R-squared based on fixed effects. Given the skew and boundedness of the FI data, we used the skew-normal distribution rather than a Gaussian model.

Supplementary Figure 3: Descriptive statistics for frailty index (FI) by assessment, burst, and sex

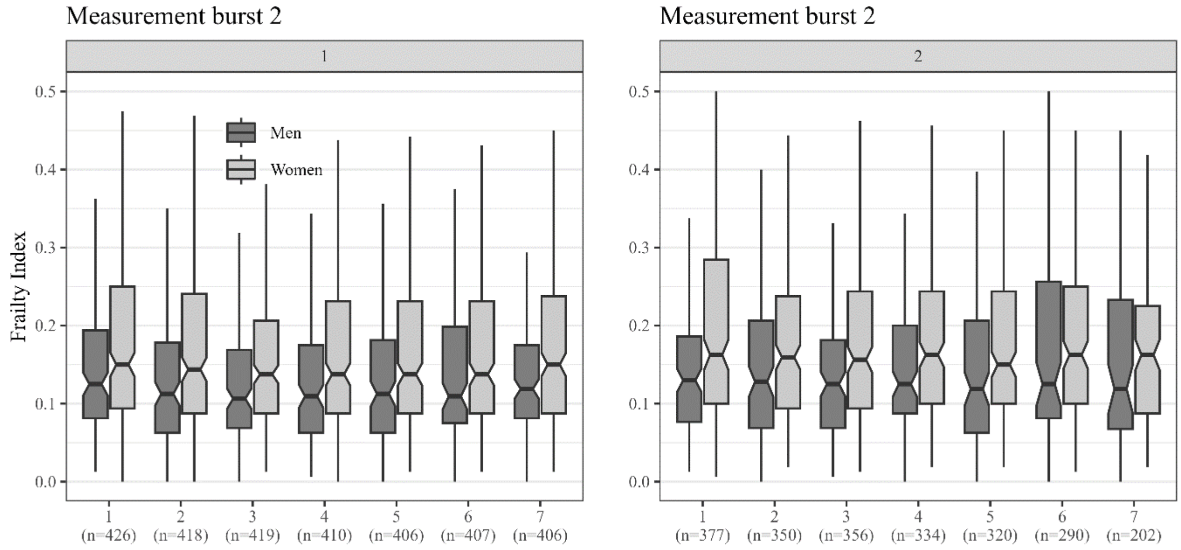

Supplementary Figure 4: Distribution of intraindividual variability (iIQR) values by health deficits of frailty index (first burst)

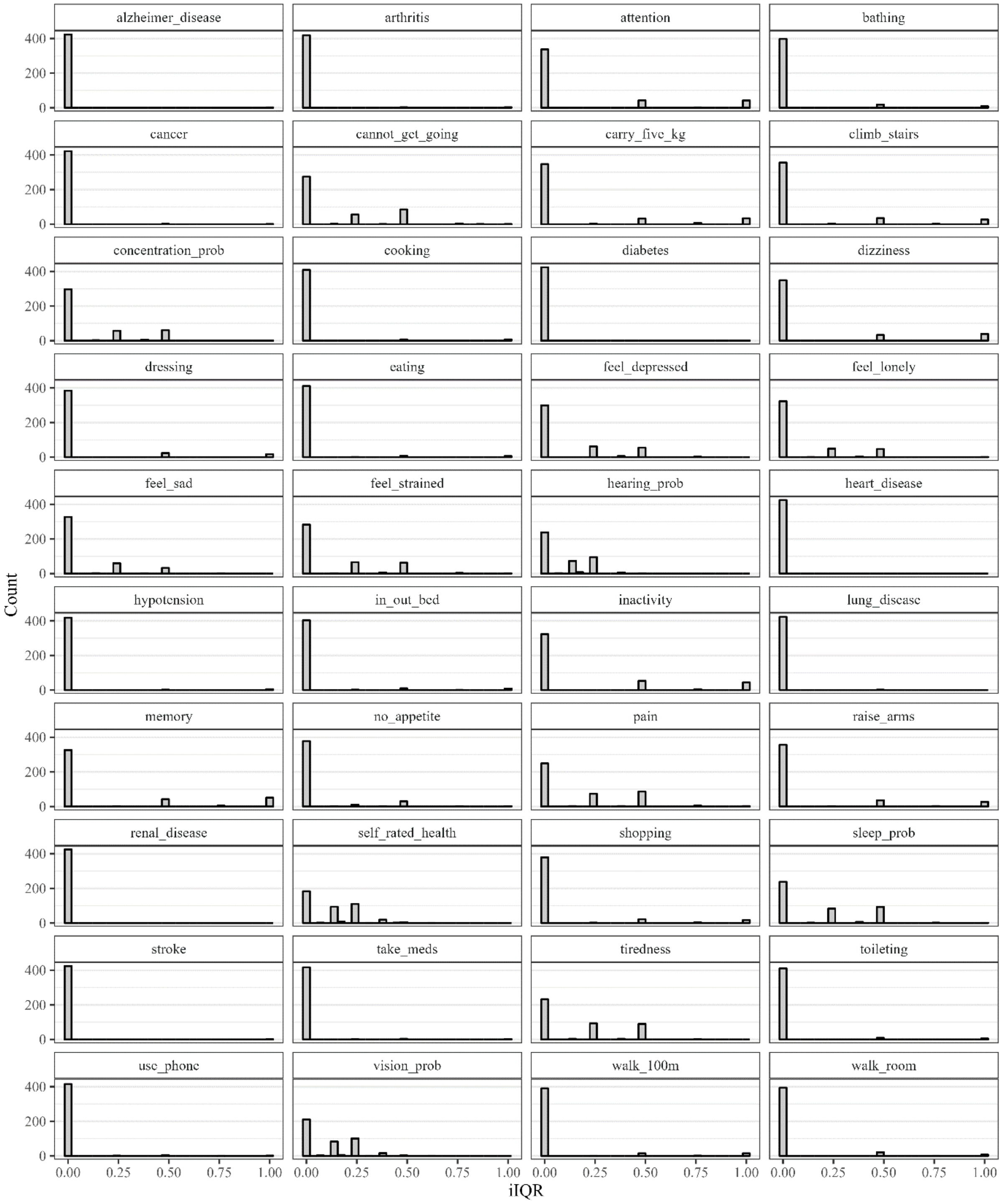

Supplementary Table 2: Short-term fluctuations at health deficit level: proportion of intraindividual interquartile range (iIQR) categories (first burst)

| | Short-term fluctuations (categories) | | |
|---|---|---|---|
| Deficit | None (IQR=0) | Some (IQR <= 0.5) | High (IQR > 0.5) |
| Self-rated health | 183 (43.1%) | 241 (56.7%) | 1 (0.2%) |
| Vision problems | 211 (49.6%) | 213 (50.1%) | 1 (0.2%) |
| Tiredness | 232 (54.6%) | 191 (44.9%) | 2 (0.5%) |
| Hearing problems | 238 (56.0%) | 187 (44.0%) | - |
| Sleep problems | 238 (56.0%) | 184 (43.3%) | 3 (0.7%) |
| Pain | 249 (58.6%) | 167 (39.2%) | 9 (2.2%) |
| Cannot get going | 275 (64.7%) | 144 (33.9%) | 6 (1.4%) |
| Everything takes effort | 283 (66.6%) | 137 (32.2%) | 5 (1.2%) |
| Concentration problem | 297 (69.9%) | 127 (29.9%) | 1 (0.2%) |
| Feel depressed | 299 (70.4%) | 123 (28.9%) | 3 (0.7%) |
| Feel lonely | 323 (76.0%) | 101 (23.8%) | 1 (0.2%) |
| Physical inactivity | 323 (76.0%) | 53 (12.5%) | 49 (11.5%) |
| Memory problem | 326 (76.7%) | 43 (10.1%) | 56 (13.2%) |
| Feel sad | 327 (76.9%) | 97 (22.8%) | 1 (0.2%) |
| Attention problem | 337 (79.2%) | 44 (10.4%) | 44 (10.4%) |
| Difficulty carry 5 kg | 347 (82.1%) | 36 (8.5%) | 42 (9.9%) |
| Dizziness | 349 (82.1%) | 35 (8.2%) | 41 (9.7%) |
| Difficulty climb stairs | 356 (83.8%) | 38 (8.9%) | 29 (6.8%) |
| Difficulty raise arms | 357 (84.0%) | 38 (8.9%) | 30 (7.1%) |
| No appetite | 378 (88.9%) | 44 (10.4%) | 3 (0.7%) |
| Difficulty shopping | 379 (89.2%) | 24 (5.6%) | 21 (4.9%) |
| Difficulty dressing | 384 (90.3%) | 24 (5.6%) | 17 (4.0%) |
| Difficulty walking 100m | 390 (91.8%) | 24 (5.6%) | 17 (4.0%) |
| Difficulty walking room | 394 (92.7%) | 16 (3.8%) | 17 (4.0%) |
| Difficulty bathing | 398 (93.6%) | 18 (4.3%) | 9 (2.1%) |
| Difficulty in/out of bed | 403 (94.8%) | 12 (2.8%) | 10 (2.4%) |
| Difficulty cooking | 409 (96.2%) | 8 (1.9%) | 8 (1.9%) |
| Difficulty eating | 411 (96.7%) | 8 (1.9%) | 6 (1.4%) |
| Difficulty toileting | 411 (96.7%) | 9 (2.1%) | 5 (1.2%) |
| Difficulty using phone | 415 (97.7%) | 7 (1.6%) | 3 (0.7%) |
| Difficulty taking meds | 417 (98.1%) | 5 (1.2%) | 2 (0.5%) |
| Hypotension | 418 (98.4%) | 2 (0.5%) | 5 (1.2%) |
| Arthritis | 419 (98.6%) | 3 (0.7%) | 3 (0.7%) |
| Cancer | 422 (99.3%) | 2 (0.5%) | 1 (0.2%) |
| Dementia | 423 (99.5%) | - | 2 (0.5%) |
| Lung disease | 423 (99.5%) | 2 (0.5)% | - |
| Diabetes | 424 (99.8%) | - | 1 (0.2%) |
| Stroke | 424 (99.8%) | - | 1 (0.2%) |
| Heart disease | 425 (100%) | - | - |
| Renal disease | 425 (100%) | - | - |

Supplementary Figure 5: Distribution of intraindividual variability (iSD) values by health deficits of frailty index (first burst) including physical performance tests among subset of participants

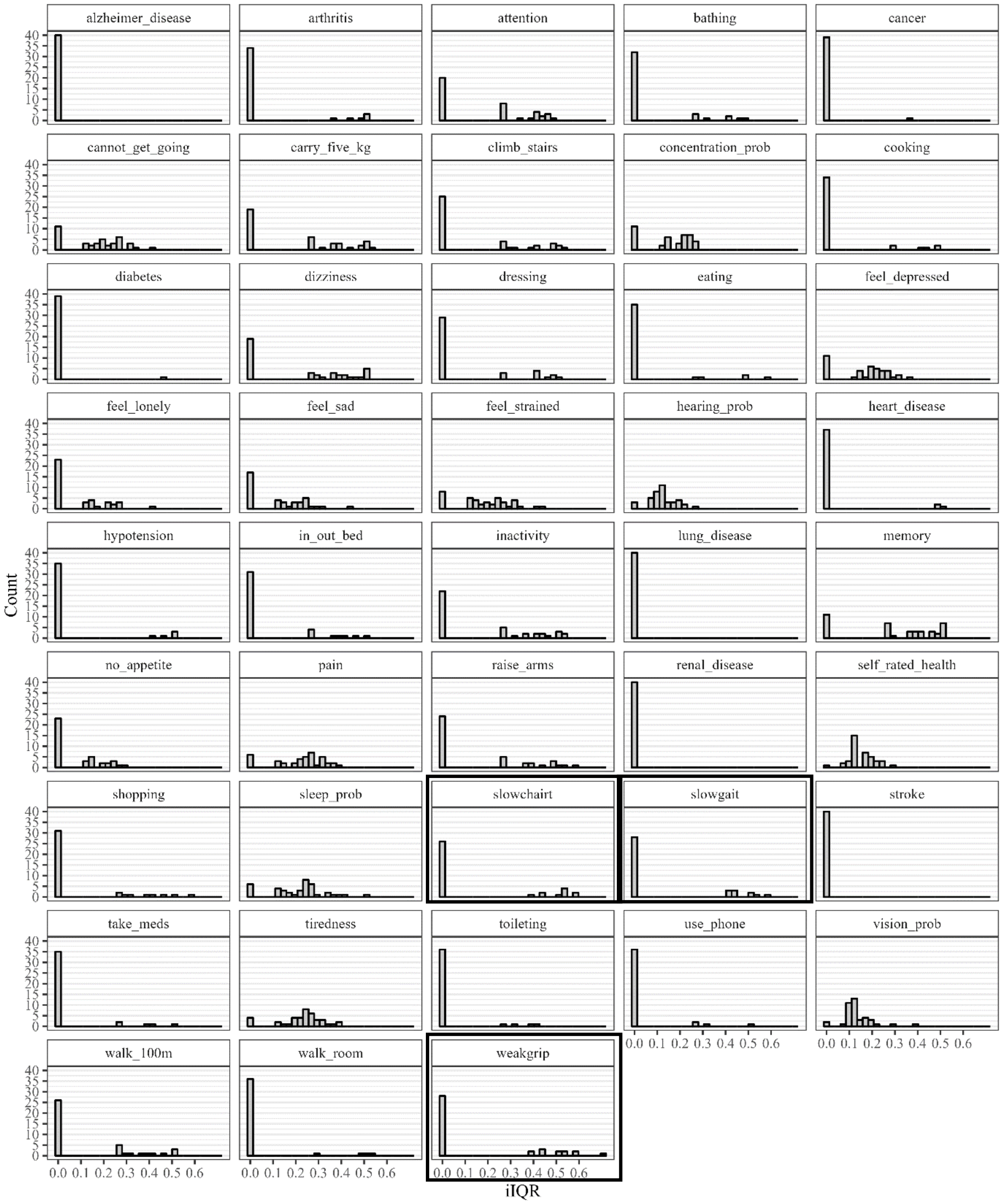

Supplementary Figure 6: Short-term frailty index fluctuations (iSD) by chronic disease status

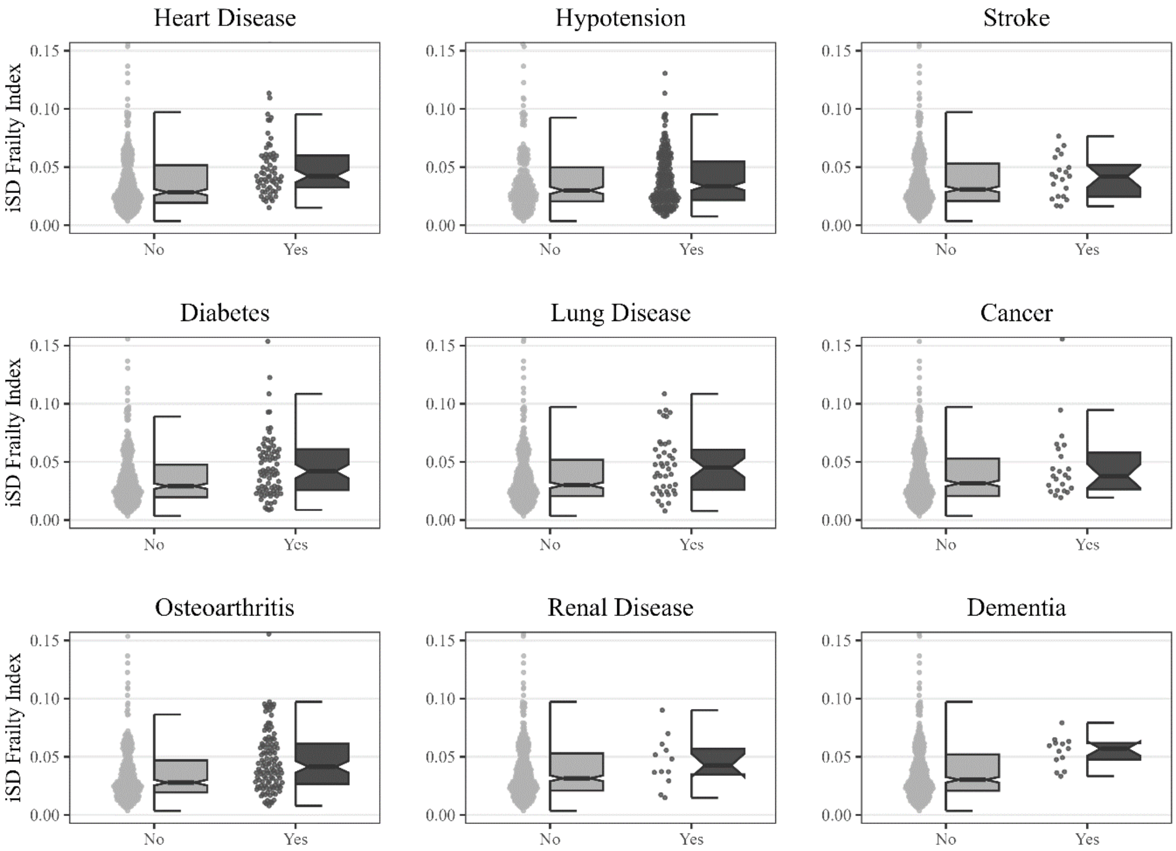

Supplementary Table 3: Results from mixed regression models

| | M0 | M1 | M2 | | M3 | | M4 | |
|---|---|---|---|---|---|---|---|---|
| | μ (95%-CI) | μ (95%-CI) | μ (95%-CI) | σ (95%-CI) | μ (95%-CI) | σ (95%-CI) | μ (95%-CI) | σ (95%-CI) |
| FIXED EFFECTS | | | | | | | | |
| Intercept | 0.19 (0.18, 0.20) | 0.18 (0.17, 0.19) | 0.18 (0.17, 0.19) | 0.04 (0.03, 0.04) | 0.28 (0.24, 0.33) | 0.05 (0.04, 0.06) | 0.27 (0.23, 0.31) | 0.04 (0.04, 0.06) |
| Wave (within-burst) | - | 0.00 (-0.00, 0.00) | -0.00 (-0.00, 0.00) | - | -0.00 (-0.00, 0.00) | - | -0.00 (-0.00, 0.00) | - |
| FI change (burst 2 vs. burst 1) | - | 0.02 (0.02, 0.03) | 0.02 (0.01, 0.02) | 0.99 (0.92, 1.06) | 0.02 (0.02, 0.03) | 0.99 (0.93, 1.07) | 0.01 (0.01, 0.02) | 0.98 (0.91, 1.05) |
| Age (years) | - | - | - | - | 0.01 (0.01, 0.01) | 1.03 (1.02, 1.04) | 0.01 (0.01, 0.01) | 1.03 (1.02, 1.04) |
| Women (vs. men) | - | - | - | - | 0.02 (-0.01, 0.04) | 1.14 (1.00, 1.30) | 0.02 (-0.01, 0.04) | 1.15 (1.02, 1.29) |
| Medium education (vs. low) | - | - | - | - | -0.04 (-0.07, -0.01) | 0.81 (0.69, 0.94) | -0.04 (-0.07, -0.01) | 0.80 (0.69, 0.92) |
| High education (vs. low) | - | - | - | - | -0.07 (-0.11, -0.04) | 0.71 (0.59, 0.84) | -0.07 (-0.10, -0.04) | 0.72 (0.61, 0.84) |
| Living alone (vs. cohabiting) | - | - | - | - | 0.02 (-0.01, 0.04) | 1.15 (1.01, 1.30) | 0.02 (-0.00, 0.04) | 1.12 (1.00, 1.26) |
| Medium social support (vs. low) | - | - | - | - | -0.08 (-0.12, -0.05) | 0.75 (0.62, 0.90) | -0.08 (-0.12, -0.05) | 0.80 (0.69, 0.92) |
| High social support (vs. low) | - | - | - | - | -0.12 (-0.15, -0.08) | 0.64 (0.53, 0.76) | -0.11 (-0.14, -0.08) | 0.68 (0.57, 0.80) |
| Dead (vs. alive in burst 2) | - | - | - | - | 0.19 (0.12, 0.26) | 1.70 (1.09, 2.55) | 0.17 (0.10, 0.24) | 1.44 (0.94, 2.13) |
| Bedrest (vs. none) | - | - | - | - | - | - | 0.04 (0.03, 0.04) | 1.50 (1.36, 1.65) |
| Fall(s) (vs. none) | - | - | - | - | - | - | 0.01 (0.00, 0.01) | 1.14 (0.97, 1.33) |
| Hospital stay (vs. none) | - | - | - | - | - | - | 0.03 (0.02, 0.06) | 1.50 (1.12, 1.97) |
| RANDOM EFFECTS | | | | | | | | |
| Individual-level intercept (SD) | 0.12 (0.11, 0.13) | 0.12 (0.11, 0.13) | 0.13 (0.12, 0.14) | 0.03 (0.02, 0.03) | 0.10 (0.09, 0.11) | 0.03 (0.02, 0.04) | 0.10 (0.09, 0.11) | 0.02 (0.02, 0.03) |
| Individual-level intercepts (COR) | - | - | 0.79 (0.73, 0.85) | | 0.69 (0.59, 0.77) | | 0.62 (0.52, 0.72) | |
| Burst-level intercept (SD) | 0.04 (0.04, 0.05) | 0.02 (0.02, 0.03) | 0.03 (0.03, 0.04) | 0.44 (0.39, 0.49) | 0.03 (0.03, 0.03) | 0.43 (0.39, 0.50) | 0.03 (0.02, 0.03) | 0.36 (0.32, 0.41) |
| Burst-level wave (SD) | - | 0.01 (0.01, 0.01) | 0.00 (0.00, 0.00) | - | 0.00 (0.00, 0.00) | - | 0.00 (0.00, 0.00) | - |
| Burst-level intercepts (COR) | - | - | 0.37 (0.21, 0.52) | | 0.37 (0.21, 0.52) | | 0.26 (0.07, 0.44) | |
| Burst-level intercept*wave (COR) | | -0.61 (-0.72, -0.48) | -0.39 (-0.59, -0.15) | 0.16 (-0.08, 0.39) | -0.35 (-0.56, -0.11) | 0.21 (-0.02, 0.43) | -0.35 (-0.56, 0.09) | 0.21 (-0.05, 0.47) |
| MODEL FIT | | | | | | | | |
| LOO | -16,552 | -17,086 | -19,225 | | -19,270 | | -19,432 | |
| R² | 0.000 | 0.006 | 0.004 | | 0.273 | | 0.293 | |

Based on 426 participants and 5,122 repeated assessments. Unweighted data. Models M1-M4 were adjusted for interview mode. μ = frailty index level, σ = frailty index fluctuations (exponentiated coefficients), 95%-CI=95% credible interval, SD=standard deviation, COR=correlation coefficient, LOO= leave-one-out cross-validation, $R^2$=Bay
